# Do Language Models Think Like Doctors?

**DOI:** 10.1101/2025.02.11.25321822

**Authors:** Liam G. McCoy, Rajiv Swamy, Nidhish Sagar, Minjia Wang, James Cao, Stephen Bacchi, Nigel Fong, Nigel CK Tan, Kevin Tan, Thomas A. Buckley, Peter Brodeur, Leo Anthony Celi, Arjun Manrai, Aloysius Humbert, Adam Rodman

## Abstract

**Background:** While large language models (LLMs) are being increasingly deployed for clinical decision support, existing evaluation methods like medical licensing exams fail to capture critical aspects of clinical reasoning including reasoning in dynamic clinical circumstances. Script Concordance Testing (SCT), a decades-old medical assessment tool, offers a nuanced way to assess how new information influences diagnostic and therapeutic decisions under uncertainty.

**Methods:** We developed a comprehensive and publicly available benchmark comprising 750 SCT questions from 10 internationally diverse medical datasets—9 previously unreleased—spanning multiple specialties and institutions. Each question presents a clinical scenario then asks how new information affects the likelihood of a diagnosis or management decision, scored against expert panels (Figure 1). We evaluated four state-of-the-art LLMs against the combined responses of 1070 medical students, 193 resident physicians, and 300 attending physicians in total across all datasets.

**Results:** LLMs demonstrated markedly lower performance on SCTs compared to their typical achievement on medical multiple choice benchmarks. GPT-4o achieved the highest performance (63.6% ± 1.2%), significantly outperforming other models (Claude-3.5 Sonnet: 58.8% ± 1.2%, o1-preview: 58.5% ± 1.3%, Gemini-1.5-Pro: 54.4% ± 1.4%). Models matched or exceeded student performance on multiple examinations, but did not reach the level of senior residents or attending physicians (Figure 2). Surprisingly, the integrated-chain-of-thought o1-preview model underperformed compared to GPT-4o, a contrast with their relative performance on other medical benchmarks.

**Conclusions:** SCT represents a challenging and distinctive benchmark for evaluating LLM clinical reasoning capabilities, revealing limitations not apparent in traditional MCQ-based assessments. This work demonstrates the value of SCT in providing a more nuanced evaluation of medical AI systems and highlights specific areas where current models may fall short in clinical reasoning tasks. We are making our benchmark publicly available in a secure format to foster collaborative improvement of clinical reasoning capabilities in LLMs.

**Brief Summary:** While large language models excel at traditional medical knowledge tests, their performance on our new public Script Concordance Test benchmark reveals important limitations in clinical reasoning capabilities, particularly in processing new information under uncertainty.

## 1. Introduction

Large language models (LLMs) have been rapidly integrated into everyday clinical care, listening to conversations between patients and their doctors, drafting patient communication, summarizing clinical care, and providing clinical decision support (CDS) at the point of care. Given persistent and growing shortages of healthcare providers in both high- and low-and-middle income countries, there has been considerable interest in using LLMs as either replacement or augmentation for clinical reasoning tasks. However, the responsible deployment of these systems requires robust, standardized evaluation frameworks that can be consistently applied as models evolve. While benchmarks alone cannot substitute for real clinical validation, carefully constructed evaluation tools that capture real-world clinical reasoning capabilities serve an essential role in the development pipeline.

Measuring the performance of LLMs on reasoning tasks is difficult and controversial. Models perform exceptionally well on medical licensing exams and question-answering data sets, but these likely reflect encoded knowledge and test-taking abilities rather than inherent reasoning^1^. Similarly, models have shown increasing performance on historical and open-ended clinical cases, such as the *New England Journal of Medicine* clinicopathological conference series^2^, but it is unclear how well these results translate to the real-life clinical reasoning capabilities of these models.

These challenges are not unique to artificial intelligence models—the assessment of reasoning has been a major goal of medical education for the teaching and assessment of human clinicians for several decades. Many measures of human reasoning have been successfully used as benchmarks of model performance, including objective structured clinical examinations with patient actors^3^, structured reflection rubrics^4^, display of reasoning^5^, and free-text multi-step clinical vignettes^6^. These evaluations have the advantage of decades of validity evidence and explicit comparisons with human clinicians, but they are also laborious to score and expensive to create, limiting their utility in large scale assessments of models, especially given persistent risks of data leakage.

Script concordance tests (SCTs) are an assessment of reasoning that is readily scalable for both human and machine evaluation. SCTs assess how new information influences both diagnostic hypotheses and management decisions, and is a proxy for an essential clinical skill – the ability to adjust clinical judgment in response to evolving information under considerable uncertainty^7^. Each question begins with a clinical vignette containing elements of natural diagnostic or management ambiguity, similar to real-world practice (Figure 1). Examinees are then presented with a potential intervention, diagnosis, or management option, followed by a new piece of clinical information. They must assess how this new information changes the likelihood of their initial hypothesis, quantified on a five-point scale from ‘Much less likely’ to ‘Much more likely.’ Answers are scored against a reference panel of experienced clinicians, with maximum credit awarded for selecting the modal response and partial credit for answers selected by at least some panel members. This scoring approach reflects the reality that even expert clinicians may reasonably disagree about the interpretation of clinical data, particularly in complex or ambiguous cases.

**FIGURE 1.**
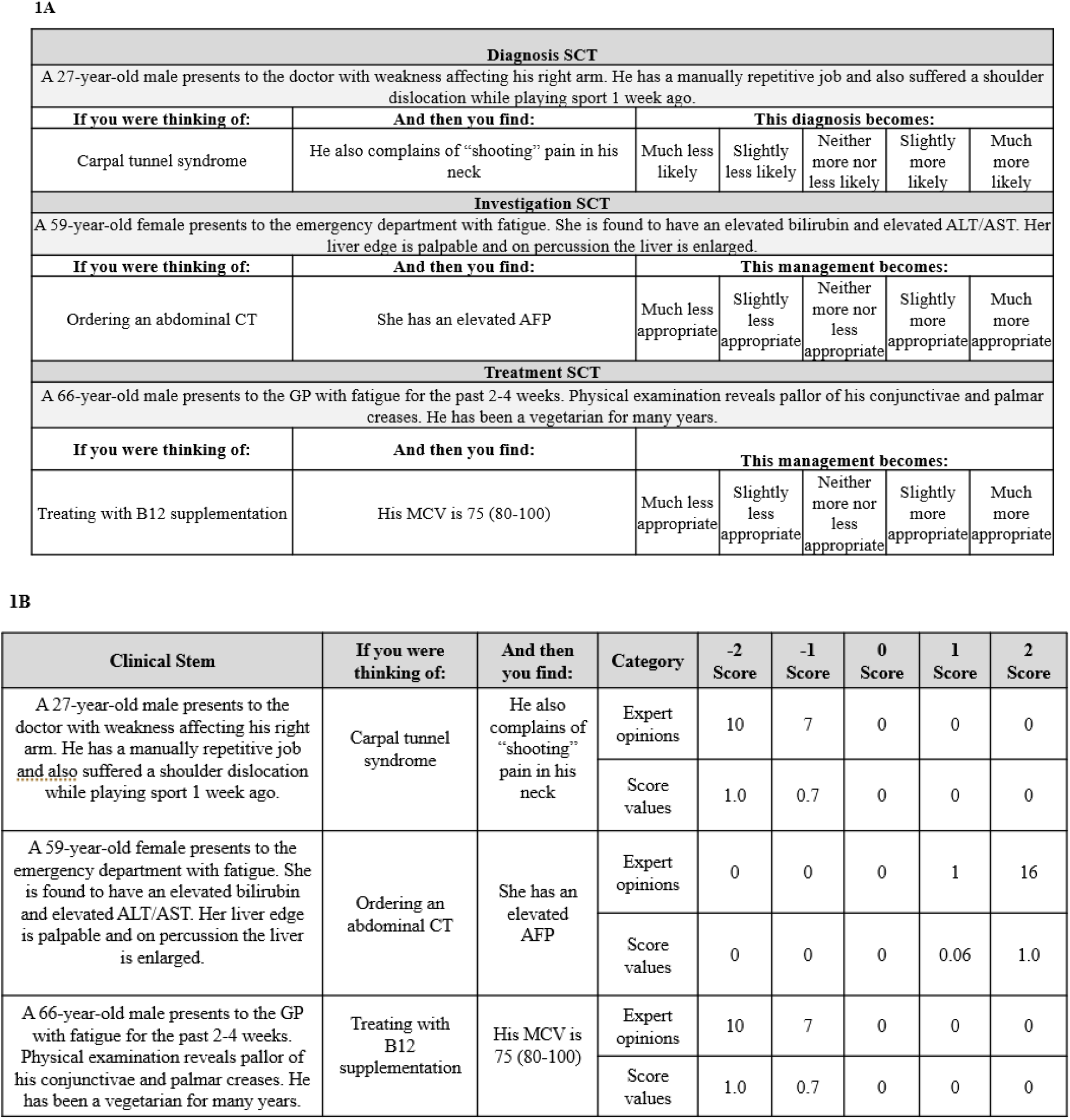
Example of a Script Concordance Test with Scoring Schema. 1A demonstrates formatting for students, 1B demonstrates formatting for LLM use, as well as an example of the scoring structure.

This paper introduces SCTs as a novel benchmark for the scalable assessment of reasoning models. Our investigation has three primary aims: first, to assess how effectively LLMs can process and integrate new clinical information to adjust their diagnostic and therapeutic reasoning; second, to compare model performance with human medical trainees and practitioners at various levels of expertise; and third, to explore how different model architectures influence performance on these specialized reasoning tasks.

By applying this established medical assessment tool to AI evaluation and making our benchmark available to the research community (https://www.concor.dance/), we not only provide new insights into the current state of LLM clinical reasoning but also establish a new methodological framework for assessing medical AI capabilities, demonstrating how validated clinical assessment tools can be adapted to provide more sophisticated evaluation of AI clinical reasoning capabilities. The SCT’s ability to reveal specific gaps between model and expert performance offers an important stepping stone between traditional benchmarks and clinical deployment, helping identify where models need improvement before being tested in real clinical settings.

## Methods

### Benchmark Development

We curated a comprehensive evaluation benchmark comprising 750 SCT questions drawn from ten distinct medical datasets. These datasets represent a diverse range of geographic locations, clinical domains, and medical specialties, as detailed in Table 1. Each SCT in our benchmark was validated through scoring by an expert reference panel following standard SCT methodology. For a subset of the tests, we additionally had access to detailed performance data from medical trainees at various levels of training, enabling direct comparison between model and human performance. Only one of these datasets, comprising 72 questions, has been previously made publicly available.

**Table 1.** Outline of Included Script Concordance Tests. ^*^Not Previously Published

| Test | Name | Source Institution | Profession | Specialty | Benchmark comparison | Detailed Response Data? | Publicly Available? | Questions (Total 750) |
| --- | --- | --- | --- | --- | --- | --- | --- | --- |
| 1 | Open Medical SCT<sup>12</sup> | University of Florida | Physician | Pediatrics | No | No | Yes | 72 |
| 2 | Adelaide SCT* | University of Adelaide | Physician | Medicine, Surgery, Psychiatry, Pediatrics, Obstetrics | No | No | No | 102 |
| 3 | Infant LP SCT<sup>16</sup> | Children's Hospital at Montefiore | Physician | Pediatrics - Infant LP | 226 Attending physicians | No | No | 32 |
| 4 | IU SCT-EM<sup>17</sup> | Indiana University | Physician | Emergency Medicine | 314 4th Year Medical Students, 40 Residents, 13 Attending Physicians | Yes | No | 59 |
| 5 | IU Student SCT<sup>18</sup> | Indiana University | Physician | Emergency Medicine | 411 2nd Year Medical Students, 70 4th Year Medical Students, 30 Attending Physicians | Yes | No | 75 |
| 6 | IU National SCT* | Indiana University | Physician | Emergency Medicine | 171 4th Year Medical Students | Yes | No | 100 |
| 7 | Neurology<br>SCT<sup>19</sup> | McGill<br>University | Physician | Neurology | 8 Medical<br>students, 41<br>Resident<br>Physicians,<br>and 16<br>Attending<br>Physicians | Yes | No | 94 |
| 8 | Physiotherapy<br>SCT<sup>20</sup> | University<br>of<br>Indianapolis | Physiotherapist | General<br>Physiotherapy | 44 First Year<br>DPT<br>Students<br>and 15<br>Experienced<br>Physiotherapists | No | No | 90 |
| 9 | Singapore<br>IM SCT<sup>21</sup> | Singapore<br>Health<br>Services | Physician | General<br>Internal<br>Medicine | 33 Interns,<br>22 Medical<br>Officers, and<br>20 Senior<br>Residents | No | No | 73 |
| 10 | Singapore<br>Neurology<br>SCT<sup>22</sup> | National<br>Neuroscience<br>Institute | Physician | Neurology | 52 Medical<br>Students,<br>37, Resident<br>Physicians,<br>and 15<br>Attending<br>Physicians | No | No | 53 |

All tests were processed through a standardization process to organize questions into a similar format (Figure 1B). Where available from test contributors (Tests 4-7, Table 1), detailed performance data was similarly standardized. For other tests (Tests 3, 8-10), performance information was extracted directly from the relevant publication.

### Model Evaluation

We evaluated four state-of-the-art large language models: OpenAI’s GPT-4o “gpt-4o-2024-08-06” (4o)^8^ and OpenAI o1-preview “o1-2024-12-17” (o1-preview)^9^, Anthropic’s Claude 3.5 Sonnet “claude-3-5-sonnet-latest” (Claude)^10^, and Google’s Gemini-1.5-Pro “gemini-1.5-pro-002” (Gemini)^11^. These models were selected as they represented the current leading edge of generally available large language models at the time of project initiation, and have demonstrated strong performance on other medical benchmarks.

The evaluation pipeline tested each model under multiple conditions: zero-shot inference, where models were given only basic instructions; few-shot inference, where models were shown five example questions with answers; and both conditions with and without prompting for explanatory reasoning. All models were used at default settings including temperature.

For few-shot inference, we selected five exemplar questions from the University of Florida public dataset^12^, with one example representing each point on the 5-point Likert scale (−2 to +2). These examples were chosen based on high consistency in expert panel scoring. The system prompt remained at default settings across all conditions, while the user prompt contained the guidelines outlining the SCT format and scoring criteria, example questions and answers in few-shot conditions, and the test question itself. Complete prompting details are provided in Supplement (Figure S1). All results are reported in line with the TRIPOD-LLM^13^ guidelines (Attached file in Supplement).

### Statistical Analyses

Primary analysis was performed on aggregate model performance across all prompting conditions. Given the non-parametric bimodal nature of SCT responses, we used a repeated-measures Friedman test to compare performance between models. Post-hoc comparisons were conducted using Conover tests with Bonferroni correction for multiple comparisons. For comparison of the overall model response distributions (Figure 4), Kruskal-Wallis with post-hoc Dunn’s test was performed. Statistical analysis performed using Scipy^14^ and scikit-learn^15^ packages.

### Benchmark Availability and Usage

To facilitate collaborative improvement of clinical reasoning capabilities in LLMs, we are making our benchmark and leaderboard available through a secure evaluation platform (https://www.concor.dance/). Two of the tests (Open Medical SCT and Adelaide SCT) will be publicly available for model fine-tuning. After completing a data use agreement to ensure appropriate usage and protect test integrity, researchers will be able to submit models to be run against the held-out test set (consisting of the remaining tests). Our private evaluation server will support use of up to 2x 80GB A100s to support open-source LLM development. Model scores will be posted to the online leaderboard comparing the best models to students and physicians.

## Results

### Overview of Test Implementation

All included models were able to successfully complete the benchmark, with parseable response rates varying between 96.2 and 99.7% (Supplemental Table 1). There was some mild variation in model performance based on the prompting strategies (Supplemental Figure S2).

### Model Performance Comparison

In aggregate analysis across all prompting conditions, GPT-4o demonstrated superior performance (63.6% ± 1.2%) (Mean ± Standard Error of the Mean (SEM)) compared to other models (Figure 2). Claude (58.8% ± 1.2%) and o1-preview (58.5% ± 1.3%) performed equivalently, with both significantly outperforming Gemini (54.4% ± 1.4%). All differences were statistically significant except between Claude and o1-preview (p > 0.05).

**Figure 2.**
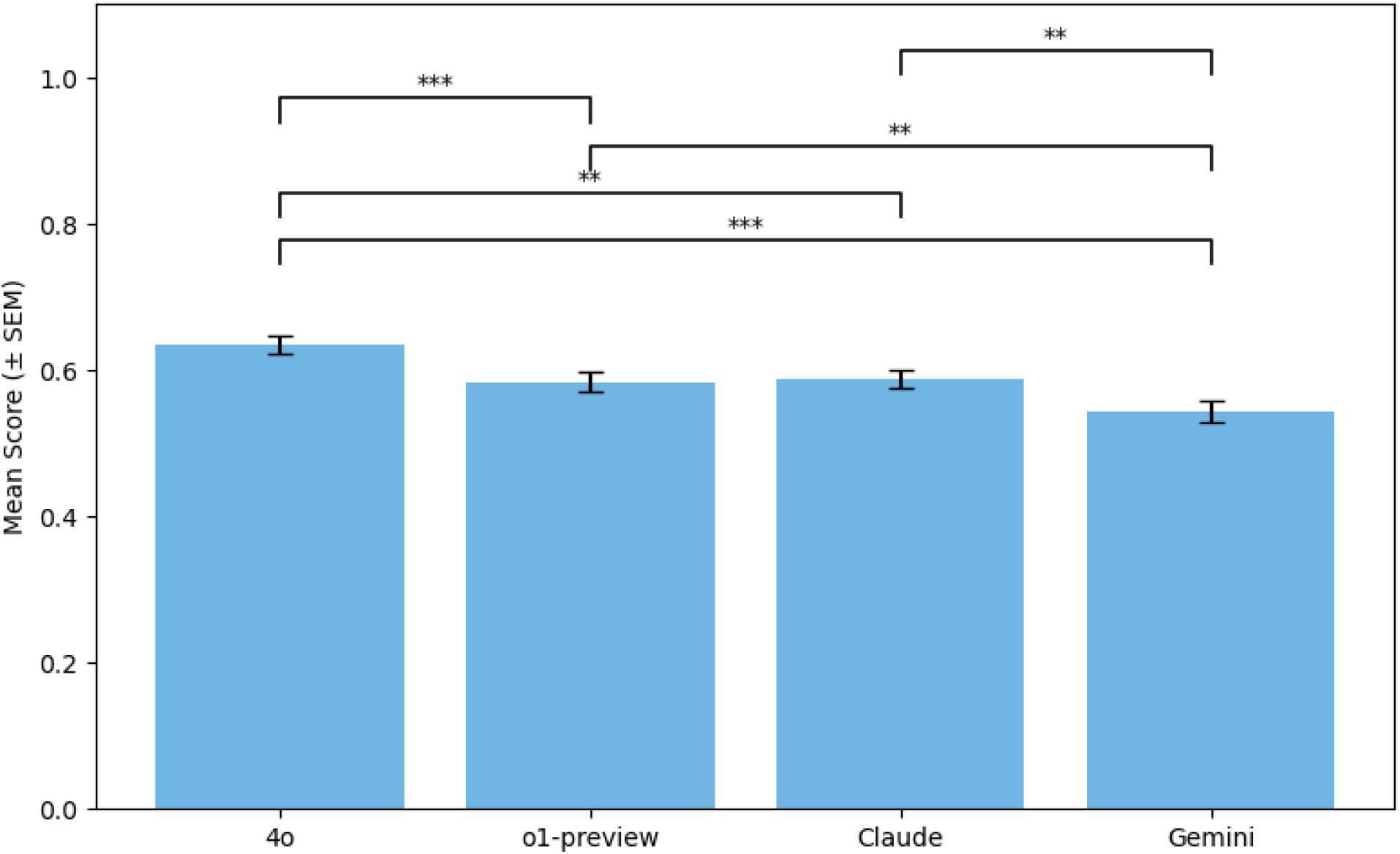
Overall Performance of Models on Full SCT Benchmark. Scores reported as mean percentage of maximum possible score, +/−standard error of mean (SEM). Statistical analysis performed with repeated measures Friedman test with post-hoc Conover test with Bonferroni correction applied. ^*^p<0.05, ^**^p<0.01, ^***^p<0.001

### Per-Test Performance

Model performance varied substantially across individual tests (Figure 3). Between all tests, comparison data was available for 300 attending physicians, 193 resident physicians, 15 staff physiotherapists, 1026 medical students, and 44 physiotherapy students. While models matched or exceeded student performance on several examinations, they consistently performed below the level of senior residents and attending physicians. All tests including multiple levels of human clinician skill show a clear positive performance gradient with expertise, underscoring the face validity of the SCT as an assessment of clinical skill.

**FIGURE 3.**
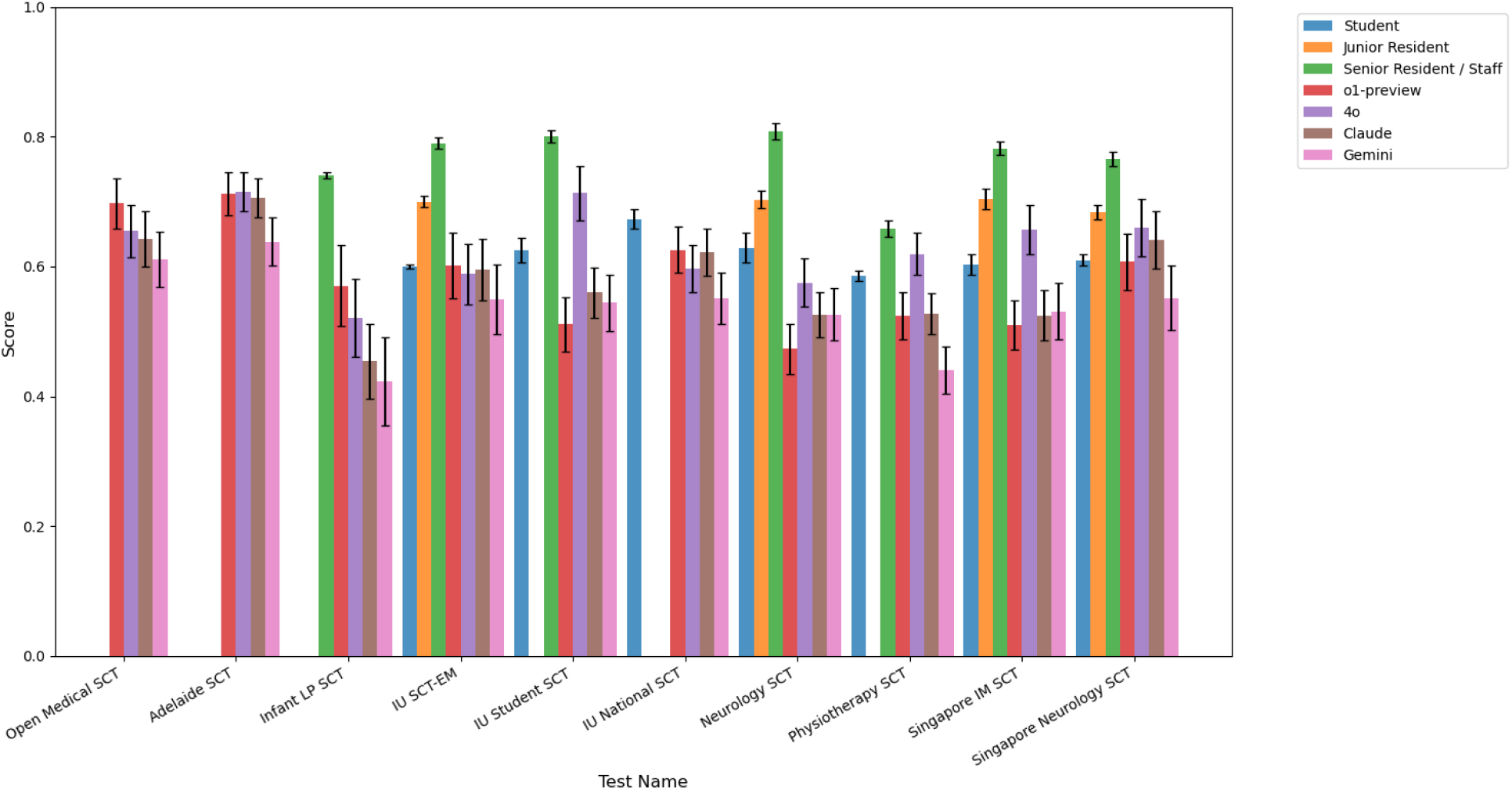
Per-Test Performance of Models and Humans of Varying Training Levels. Results shown as the mean ratio relative to the maximum possible score, +/−SEM.

**FIGURE 4.**
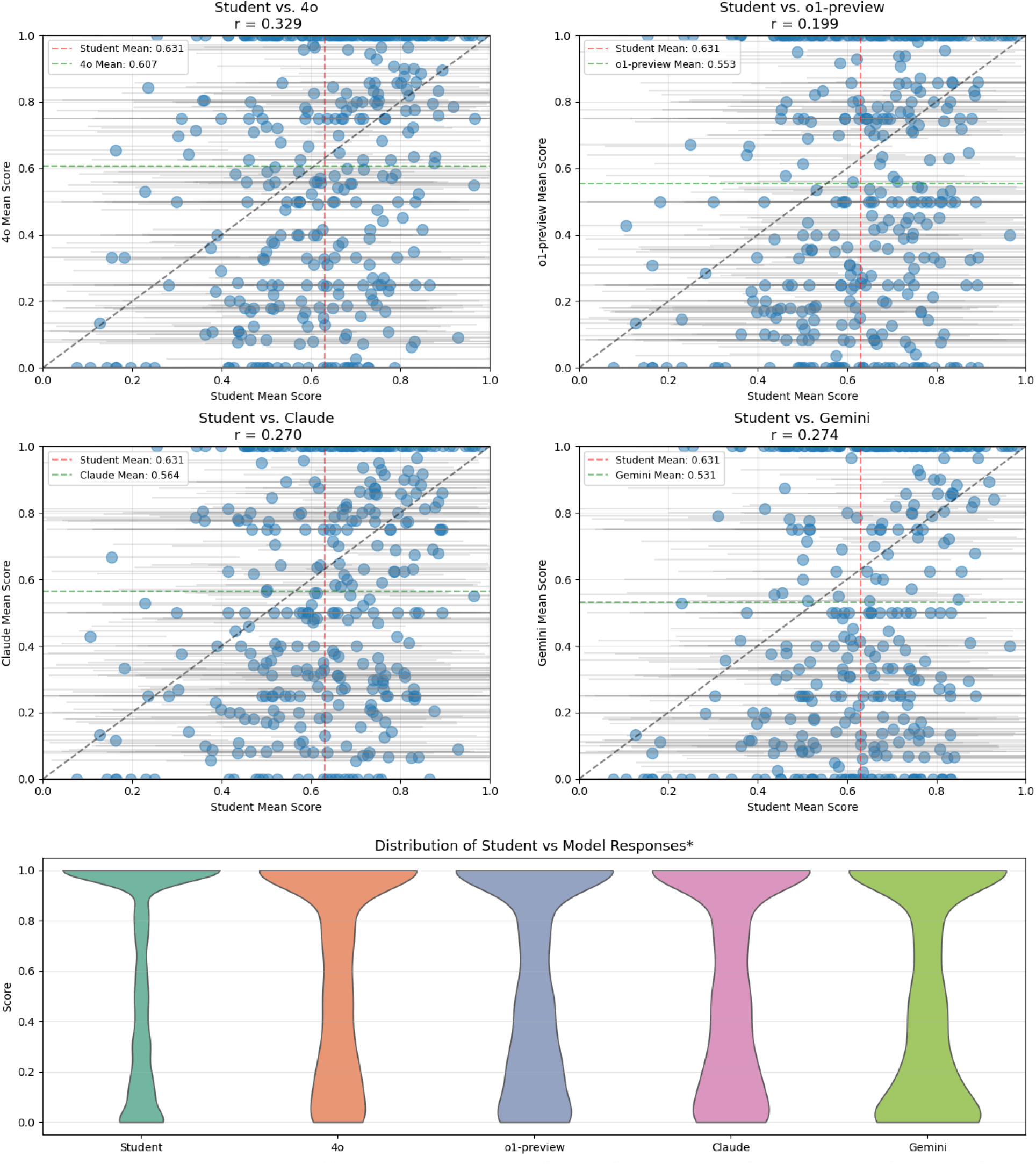
Detailed Comparison of Student and Model Responses. Comparison performed on all three IU Datasets and Neurology dataset where detailed student response data are available. Average student performance and average model performance per question is shown in scatter plots, while violin plots encompass all student and model answers. ^*^Comparison of distributions performed with Kruskal-Wallis test (H = 430.884, p < 0.001), and post-hoc Dunn’s test (all pairwise comparisons significant p < 0.0001 except o1-preview vs Claude (p = 1.00 post-adjustment)).

### Detailed Comparison with Student Responses

As shown in Figure 4, we perform detailed comparison between model and student responses where available (for all three Indiana University Emergency Medicine tests, and the McGill Neurology test). We see overall a weak correlation between the difficulty of questions for students and the models, with the correlation for o1-preview particularly poor. Many questions that are difficult for students are answered easily by models, and vice versa. With reference to the violin plots, we see that student responses particularly cluster around perfect scores, while the models demonstrate a more rounded curve. We see that the models are all significantly different from the student response distributions, as well as from each other (except for o1-preview and Claude).

## Discussion

Our study demonstrates that Script Concordance Tests represent a distinctive and challenging benchmark for evaluating clinical reasoning in Large Language Models. Unlike many existing medical benchmarks where LLMs have achieved superhuman performance^2^, SCTs reveal significant gaps between model and expert clinician performance. This finding is particularly notable given recent concerns about benchmark saturation^23^ in medical AI evaluation. In making this benchmark publicly available, we hope to enable novel work for the improvement of LLM reasoning both clinically and in more general domains.

The pattern of model performance on SCTs provides important insights into the nature of LLM clinical reasoning capabilities. While models can match or exceed medical student performance on many questions, they consistently fall short of expert clinician levels—a pattern that differs markedly from their performance on traditional multiple choice questions (MCQs). This discordance between SCT and MCQ performance is not unique to AI systems; studies of emergency medicine residents have found no significant correlation between SCT and MCQ scores^24^, suggesting these tests evaluate fundamentally different aspects of clinical capability. The distinction becomes more pronounced with increasing expertise—while medical students’ early performance shows correlation between MCQs and SCTs, this correlation disappears in later years of training^25^. This pattern suggests that as clinicians develop more sophisticated clinical reasoning skills, their performance on SCTs begins to reflect capabilities beyond simple biomedical knowledge. Given robust evidence of knowledge encoding in models, increased gains of performance on SCTs may suggest improved reasoning abilities.

The relative performance of different model architectures on our benchmark yields additional insights into the nature of LLM clinical reasoning. The underperformance of the integrated chain-of-thought o1-preview model compared to GPT-4o is particularly striking and unexpected, given that o1-preview typically outperforms GPT-4o substantially on most benchmarks^2,9^, including medical ones. This represents, to our knowledge, the first medical benchmark to invert this pattern. This outperformance is not universal on a per-test or per-question basis (Figure 3, Supplemental Figure 3), and the reasons for this disparity require further investigation.

These findings underscore a broader challenge in evaluating clinical reasoning: the most epistemologically sound measurements are those validated against expert human performance. While high clinician performance on standardized assessments correlates with better patient outcomes^26^, such measurements alone are insufficient for evaluating clinical competence. Over the past several decades, medical education has evolved to a competency-based assessment system, in which tasks are evaluated in actual clinical care by expert supervisors^27^. Competency-based medical education (CBME) lacks robust evidence of improvement in patient outcomes but it does stress the importance of broader – and often subjective – measures of performance. Such evaluations cannot clearly be performed in automated fashion by computerized algorithms.

Recent efforts to address these limitations have led to the development of agent-based benchmarks that attempt to simulate complete clinical encounters through LLM-based role-playing^28,29^. However, these simulations, while technologically sophisticated, may further abstract away from the concrete realities of clinical decision-making. Our findings suggest a more grounded path forward: continuing to develop human-validated benchmarks that target specific, well-defined aspects of clinical reasoning, while acknowledging that these tools will eventually face their own limitations. Ultimately, just as medical education has evolved to emphasize direct observation of clinical performance, the true test of LLM clinical reasoning capabilities will require careful evaluation through controlled clinical trials—a process that our work helps lay the groundwork for by identifying specific areas where current models may require additional development before deployment.

### Strengths

Our benchmark has several key strengths. First, it includes SCTs from diverse geographic and practice contexts, spanning North America, Asia, and Australia, and encompassing both general and specialty practice settings. This diversity is particularly important given that clinical medicine varies substantially by region and context—differences in disease prevalence, resource availability, and standard practices can all impact clinical decision-making. The strong performance correlation across these varied settings suggests that the models’ clinical reasoning capabilities are robust across different practice environments, though their performance consistently falls short of expert clinicians. Second, it includes human performance benchmarks by which to compare performance; several of the SCTs also contain the responses from multiple training levels, showing stepwise improvement in performance with additional experience. Finally, for all but one of the question sets, there is no risk of data leakage as the questions have never been previously published.

### Limitations

Several important limitations must be considered when interpreting these results. First, as with all LLM evaluations, performance on the SCT must be regarded as highly prompt- and context-dependent. These results should be taken as a proof of concept for the difficulty of the SCT, but they do not necessarily establish a ceiling for the included LLMs’ performance. Advanced prompting strategies^30^, explicit Bayesian reasoning approaches, or tool use may improve performance beyond what we demonstrate here.

Second, there are inherent limitations to the SCT methodology itself that warrant consideration. These include:

1. Response distribution bias: Reference panel members select extreme options less frequently than novice test takers in certain tests^31^, creating a potential scoring artifact where avoiding extreme responses could artificially inflate scores regardless of clinical reasoning ability.
2. Aggregate scoring challenges: The SCT’s assumption that all reference panel opinions have validity can lead to factually incorrect responses receiving partial credit^32^.
3. Scale interpretation ambiguity: There may be inconsistent interpretation of the Likert scale between examinees and experts, with some viewing the extremes as representing confidence levels rather than magnitude of probability adjustment^33^. This ambiguity could affect both human and AI performance metrics.

Third, the benchmark’s geographic and institutional diversity, while a strength, also introduces potential confounders. Context specificity is an increasingly recognized problem in the evaluation of reasoning^34^, and the validity of using reference panels from one healthcare context to evaluate performance in another remains unclear. While our results suggest robust performance across different settings, this may not generalize to all clinical contexts or decision-making scenarios. Approaches such as the SCT are useful for broad benchmarking, but examination-based benchmarks are insufficient for the forms of evaluation which are necessary to enable meaningful real-world deployment^35^.

Fourth, our evaluation methodology focused on aggregate performance across prompting strategies. While this provides a robust overall assessment, it may mask important variations in model performance under specific conditions or with particular prompting approaches. Future work should investigate these variations more systematically and explore how different prompting strategies might better align with the cognitive processes SCTs aim to evaluate.

Finally, our study – and all concordance-based evaluation in the LLM performance – demonstrates a fundamental challenge in assessing AI systems as they approach human-level performance: how do we evaluate performance when there is no clear “ground truth”? In clinical medicine, expert physicians frequently disagree on optimal approaches, particularly in complex or ambiguous cases. The SCT’s scoring methodology, which awards partial credit based on the distribution of expert responses, explicitly acknowledges this reality of clinical practice. However, this approach raises important questions as AI systems begin to match or exceed human performance in specific domains. It is conceivable that a superhuman AI may move (correctly) away from the human consensus on certain clinical decisions, yet such performance would harm its score on the SCT.

These limitations do not invalidate our findings but rather highlight important considerations for future research and development in both medical AI evaluation and SCT methodology. They also emphasize the need for multiple, complementary approaches to assessing clinical reasoning capabilities in AI systems.

## CONCLUSION

Our SCT benchmark, now publicly available to the research community, reveals important limitations in LLM clinical reasoning capabilities that are not apparent in traditional benchmarks. While current models can match or exceed medical student performance on many SCTs, they consistently fall short of senior resident and expert clinician performance, particularly in cases requiring nuanced interpretation of ambiguous clinical data. The unexpected performance patterns observed among models—particularly GPT-4o’s superior performance compared to the typically stronger o1-preview model—suggest that current approaches to enhancing LLM reasoning capabilities may not fully address the requirements of clinical decision-making.

Our results highlight broader challenges in evaluating AI systems as they approach human-level performance in medicine, particularly in domains where expert disagreement is both common and clinically appropriate. The SCT’s concordance-based scoring system, which acknowledges legitimate disagreement among experts, offers a more sophisticated framework for assessing clinical reasoning than traditional right/wrong evaluations. However, this approach also raises important questions about how to evaluate AI systems that may potentially exceed human consensus in certain areas, particularly as these systems move closer to clinical deployment.

To facilitate ongoing development, we are making portions of our benchmark publicly available for model development, with the remainder accessible through a secure evaluation framework. This tiered access approach balances the need for open science and reproducibility with maintaining test integrity. By providing both training data and held-out test sets, we enable researchers to develop and validate new approaches while ensuring meaningful comparisons across different models and methodologies. As the field of medical AI continues to evolve rapidly, maintaining such standardized evaluation frameworks—particularly those validated against human expert performance—will be crucial not only for tracking technical progress, but ultimately for ensuring that AI systems can safely and effectively support clinical decision-making in real-world healthcare settings.

## Data Availability

Two of the ten included benchmark datasets are publicly available. To protect test set integrity, the remaining data in the benchmark is available for public submission of models for testing (https://concor.dance).

## Ethics Review

In all cases where detailed student data was used, it is covered by the original institutional review boards for the component datasets. For IU SCTs #4-6, the institutional review board of the Indiana University School of Medicine (IUSM) gave ethical approval. For Neurology SCT #7, the McGill University Institutional Ethics Board gave ethical approval. For all other SCTs, any human performance data used in this paper was extracted in summary form from the openly published final papers as cited in **Table 1**, which were open to the public prior to the initiation of the study.

## Data and Code Availability

Given the privacy agreements with the various research groups who have agreed to provide these tests, data access will be available in the form of a hosted benchmark available for model testing upon signature of a data use agreement (https://concor.dance).

## Acknowledgements

We thank the authors of the script concordance tests who graciously agreed to provide us with access to their prior work. In addition to the test creators who are co-authors on this paper, we wish to acknowledge Stuart Lubarsky, Lindsey Kojich, Stephanie A. Miller, Todd P Chang, Martin V Pusic, Nnenna O Chime, Peggy Seidman, Mary Johnson, Bart Besinger, and Pranshu Bhardwaj.

